# Estimation of Near-kink Reproduction Numbers During the Emergent Variants of the COVID-19 Pandemic: Log-quadratic and Forward-imputation Approach

**DOI:** 10.1101/2023.04.24.23289029

**Authors:** Ichiro Nakamoto

## Abstract

**Background:** Sketching the major portraits of the COVID-19 epidemic when variants of the pathogen emerge is critical to inform the dynamics of disease transmission, reproduction (i.e., the average counts of individuals of secondary infections generated by an index individual infected by the virus) strength of the pathogen, and countermeasure strategies. Multiple approaches, including log-linear, EpiEstim (an R package generally utilized to estimate the evolution traits of epidemics), and near-log-linear techniques, have been exploited to evaluate the principal parameters such as basic and effective reproduction numbers of an epidemic outbreak.

**Objective:** This study focuses on the kink corner (i.e., sharp alternation of direction of the transmission curve) presenting differentiated log-quadratic traits where more infectious variants of viruses emerge at the diminishing transmission phase of an infectious disease.

**Methods:** A novel log-quadratic trending framework was proposed to project potentially unidentified cases (i.e., forward imputing approximately one week ahead) of COVID-19 around the kink, where the transmission of the pandemic initially lowered and accelerated subsequently, and exercised with the updated framework of classic EpiEstim and Log-linear model. I first compared the performance near the kink using the proposed technique versus the two traditional models taking into account a variety of levels of transmissibility, data distribution (Weibull, Gamma, and Lognormal distributions), and reporting rates (0.2, 0.4, 0.6, 0.8 and 1.0 respectively). Thereafter I utilized the revised framework on the outbreak data of four settings including Bulgaria, Japan, Poland, and South Korea from June to August 2022.

**Results:** The proposed framework reduced the estimation bias versus traditional EpiEstim and log-linear methods near the kink. The coverage estimates of 95% confidence intervals improved. The proposed forward-imputation method implied generally a consistent ascending trend of effective reproduction number estimation applying to a precipitous transition from diminishing to diverging scenarios versus the irregular zigzagging outcomes in classic methods when more contagious variants of the virus were present in the absence of effective vaccines.

**Conclusions:** The log-quadratic correction accounting for transmissibility, data distribution, reporting rates, sliding windows, and generation intervals improved the basic and effective reproduction numbers estimation at the kink corner versus the classic EpiEstim and log-linear models by refined amendment of curve fitting. This is of concern when essentially the fundamental transmission traits of a pandemic alter expeditiously and countermeasures are needed at the earlier variant phases of the transiting climax with the advancement of the pandemic.

## Introduction

The COVID-19 pandemic witnessed multiple waves of variants such as alpha, beta, delta, and omicron since the outbreak was officially established in late 2019 [1]. Findings reveal that the effectiveness of vaccines is compromised on average by existing variants of COVID-19 [1,2]. These variants have introduced significant challenges in curtailing the transmission of the pathogen and the treatment of infections. The fundamental traits of mutations include intensification of virus replication and thus excessive transmissibility identified recently by Delta and Omicron [3]. The latter was partially or completely resistant to neutralization and escaped most therapeutic monoclonal antibodies. Sera from recipients of the Pfizer or AstraZeneca vaccine > 5 months after vaccination scarcely inhibited Omicron [4,5]. Multiple sublineages of SARS-CoV-2 Omicron have been reported worldwide. These variants were more infectious than their previous counterparts and were known to spread rapidly [5,6].

Attributable to factors such as limited testing capacities, compromised service quality of healthcare, vaccination hesitancy, substantial pre-symptomatic or asymptomatic infections, and change of control strategies, some settings have recognized the dilemma in connection with undetected and under-reported infections [6,7,8]. For instance, the change in relaxing public health suspension of vaccine passes, and social measures were found to associate with the increased transmissibility of the Omicron variant in South Korea [7]. Japan likewise experienced a high level of COVID-19 infections with an average of nearly 200 thousand new infections per day nationwide during a short period from July to August 2022 [8]. Some countries had other problems such as vaccine hesitancy, for instance, Bulgaria reported a hesitancy level of 62% and Poland reported a similar issue in older age cohorts [9]. On the other hand, available data by July 2022 suggest that variants of COVID-19 specifically the BA.2, BA.4, and BA.5 subvariants escape neutralizing antibodies to a significant extent, which complicated the transmission of the pandemic [10-17].

The series of mutations of the pathogen usually is associated with idiosyncratic traits of the epidemic curve: a kink that potentially alters considerably the direction of the trajectory. Evaluation of the transmission potential and reproduction numbers near the kink corner is meaningful for signaling the potential of disease infection and the subsequent strategies of responses [11-20]. Hence, one critical problem remains to be the concern: how do the principal parameters (e.g., basic and effective reproduction numbers) regulating the trajectory of the pandemic behave close to the turning kink? It is of necessity that the fundamental traits of the kink are captured to reflect the transmission dynamics of the pandemic by using available data in the early phases of transition when the healthcare system, the preparedness of the public or the countermeasures are not in the status of equilibrium [21]. Basic (*R*_0_) and effective (*R*_*t*_) reproduction numbers are two fundamental metrics to measure the infectiousness of the virus transmitted from one individual to another. In this context, I revisited the fundamental concepts of basic and effective reproduction numbers respectively, and conceived a novel log-quadratic framework. Conceptually basic reproduction number *R*_0_ is defined merely at the onset of a pandemic when all individuals are completely susceptible, representing the average cases of secondary infections that an index infected individual generates. The transmission of disease holds if *R*_0_ >1 and ceases if *R*_0_ <1 [11-14]. In contrast, effective reproduction number *R*_*t*_ denotes the average cases of secondary infections caused by a single infectious individual at a given time *t* [15-17]. These two parameters potentially reflect critical information inherent in the pandemic, including the infectiousness of the pathogen, the strength of social measures, the behavior changes of individuals, the environmental factors, and the crude threshold of herd immunity [13-20], and are indispensable to proposing subsequent countermeasures [14-24].

A variety types of techniques have been employed to investigate these questions. [Luis Alvarez](https://www.pnas.org/doi/suppl/10.1073/pnas.2105112118) et al. [15] appraised *R*_*t*_ by inverting the renewal equation in connection with the observed incidence curve. Raúl et al. [16] utilized a fitting model for *R*_0_ with statistical distributions and a Bayesian approach relying on prior information to project posterior distributions. Jie Y et al. [17] used log-transformation of the daily infections to infer the impact of COVID-19. Cori et al [18] developed the EpiEstim framework and it has been known as an effective technique to estimate the instantaneous reproduction number *R*_*t*_. However, EpiEstim and other statistical models suffer from systematic bias in the early stages of an epidemic, and whether the performance can be generalized to the kink in association with variants of infectious diseases is not known [18,19]. By extending the log-linear and EpiEstim concepts and their subsequent refined frameworks [19, 20], I propose a novel method by accounting for potentially incorrect or missing information about the outbreak near the kink corner and utilizing transformed simulation data to scrutinize the effectiveness versus the other two classic models. I evaluated how the novel framework entails a reduction in the estimation bias of the basic reproduction number close to the kink in comparison with traditional EpiEstim and log-linear models. Lastly, I appraised the estimates produced by the proposed model on the early-variant transition stage outbreak data of four representative countries, where kink traits of disease transmission were observed.

## Methods

### Classic EpiEstim Model

The effective reproduction number, *R*_*t*_, can be estimated by the ratio of the cases of new infections *I*_*t*_ generated at time step *t*, to the total infectiousness of infected individuals at time *t* that is governed by 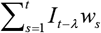, where *w*_*s*_ is the infectiveness of infected individuals [20]. Consequently, if *t* represents a time point, the form of *R*_*t*_ is transformed to the instantaneous reproduction number at a specific time point; in contrast, if *t* denotes an interval of period, the definition is converted to a sliding window portraying relatively stable transmission of the pathogen within that time slot. I focus on the period from June to August 2022 and four settings including Bulgaria, Japan, Poland, and South Korea. Studies have identified that methods including EpiEstim overestimate reproduction numbers during the preliminary stages of epidemic trending and the magnitude of bias increases with the degree of data missing. This can potentially engender misleading conclusions about the transmission dynamics of the epidemic and the subsequent countermeasures of social efforts [12-24].

### Kink and Log-quadratic Transformation

The new framework assumes that the transmission of COVID-19 follows a log-quadratic polynomial form attributable to the emergence of novel variants of the pathogen, the major traits of which are characterized by the kink remarkably switching the direction of the transmission. The log-quadratic mathematical model is asymptotically given by log(*cases*) = *a* * *day* ^2^ + *b* * *day* + *c* where *a,b*, and *c* denote respectively the day-squared, day-term, and constant coefficients of the quadratic form. And for generalization assume *a* > 0 and *b* < 0. Hence the kink of the preceding equation is determined by 2*a* * *day* + *b* = 0, that is to say, at the time 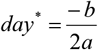 the minimum cases of log-transformation are derived as 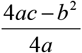. If the time satisfies 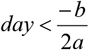 the daily increment of infections slows down and the pandemic tends to eradicate ultimately, in contrast, if the time satisfies 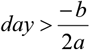, the daily increment of infections increases.

### Proposal: Correction of EpiEstim and Log-linear Framework

I propose an updated calibration to account for potentially incorrect statistics of infections around the kink corner where more infectious variants occur. Specifically, I suppose that the epidemic follows a log-quadratic distribution close to the kink to forward-impute the subsequent infections. As the transmission is generally rapid, which is reflected in the change of reproduction numbers and consequently I assume that the period of forward-imputation is not more than 7 days. Our framework can be implemented by complying with the mechanism illustrated in Figure 1. Initially, I apply a log-quadratic model on the transformed simulation observations to attain the growth rate. Then, I utilize the log-quadratic structure to forward-impute subsequent infections one week ahead, for which inaccuracy of data reporting occurs partially attributable to factors such as the low capacity of testing, ineffectiveness of vaccines, vaccination hesitancy and considerable sub-clinical infections at the early-stage transition occurrence of variants. Thereafter I follow the general schema depicted in [20] to identify the generation interval (conceptually defined as the infection time between the infector and the infected.) necessary for the forecast. I include the forward-imputation cases in the final sample and run the log-quadratic model with the EpiEstim framework to appraise the updated performance.

**Figure 1.**
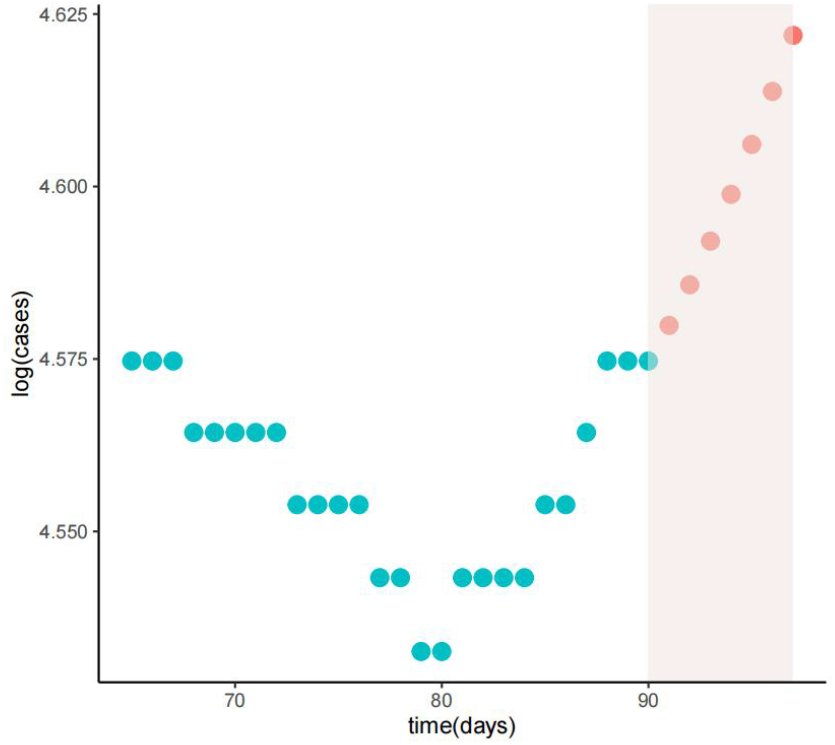
Illustration of the log-quadratic trending calibration appraised with simulation data. Logarithm transformation of the initial dots (cyan points) is utilized to fit a log-quadratic model. The novel quadratic model is then utilized to forward-impute cases (orange dots) to compensate for the incorrectly reported or missing data beyond that point during the early stage of the variants transition. Simulation parameters: simulation times are set to 500, the reporting rate *λ* of infections is 1.0, the time slot of forward-imputation is 7 days, and the window of sliding is 3 days.

### Design of Simulations

I appraised the outcome of the proposed novel framework versus traditional EpiEstim and Log-linear methods. As the region near the kink corner incorporated the key points mostly altering the transmission from convergence to divergence, I assessed *R*_0_ that was close to threshold value one by taking values of 0.9, 0.95, 1, and 1.2 with 50 simulations respectively. These values potentially capture the principal traits of the transmission. The number of individuals was equal to 1e8 and the epidemic was initiated by 5 initial cases by assumption. The average time exhausted in the exposed and infectious compartments was supposed to be 3 and 4 days respectively, from which generation interval was calculated [6, 20]. I follow the two types of imperfect reporting sketched in [20], assuming the kink points approximately distribute at day 80 in the simulation data. A variety level of under-reporting rates ranging from 0, 0.2, 0.6, and 0.8 is utilized, which corresponds to the differentiated preparedness of the healthcare system and the population. I assume the observations followed binomial distributions with the likelihood of correct reporting identical to the reporting rate divided by five.

### Ethical considerations

None applicable.

## Results

### Performance Comparison of Different Frameworks

I apply the log-linear, EpiEstim, and proposed framework to the simulation data assuming a Weibull distribution of generation interval. For other distributions including Gamma and Lognormal, I obtained similar outcomes (Supplementary Figures S18-S21). Figure 2 compares the effect of three different models around the kink and the true values of reproduction number are set to equal to 0.9, 0.95, 1, and 1.2 respectively. I set the reporting rate *λ* as 0.8. The result indicates that in all scenarios with kink, the log-linear model underestimates the values of the basic reproduction number. These trends are evaluated for other types of *λ* (Supplementary Figures S10-S17), implying consistency in the estimates obtained with different reporting rates. On the other hand, the EpiEstim framework overestimates the basic reproduction number close to the kink when *R*_0_ < 1 and underestimates the value when *R*_0_ > 1. Our adjustment enhances the range accuracy of *R*_0_ by improving the estimation of the confidence interval and performs effectively at threshold one. The average widths of 95% confidence interval are illustrated in Figure 3. Under different scenarios, the range estimation of the true reproduction number is increased using the proposed model versus the log-linear and EpiEstim models.

**Figure 2.**
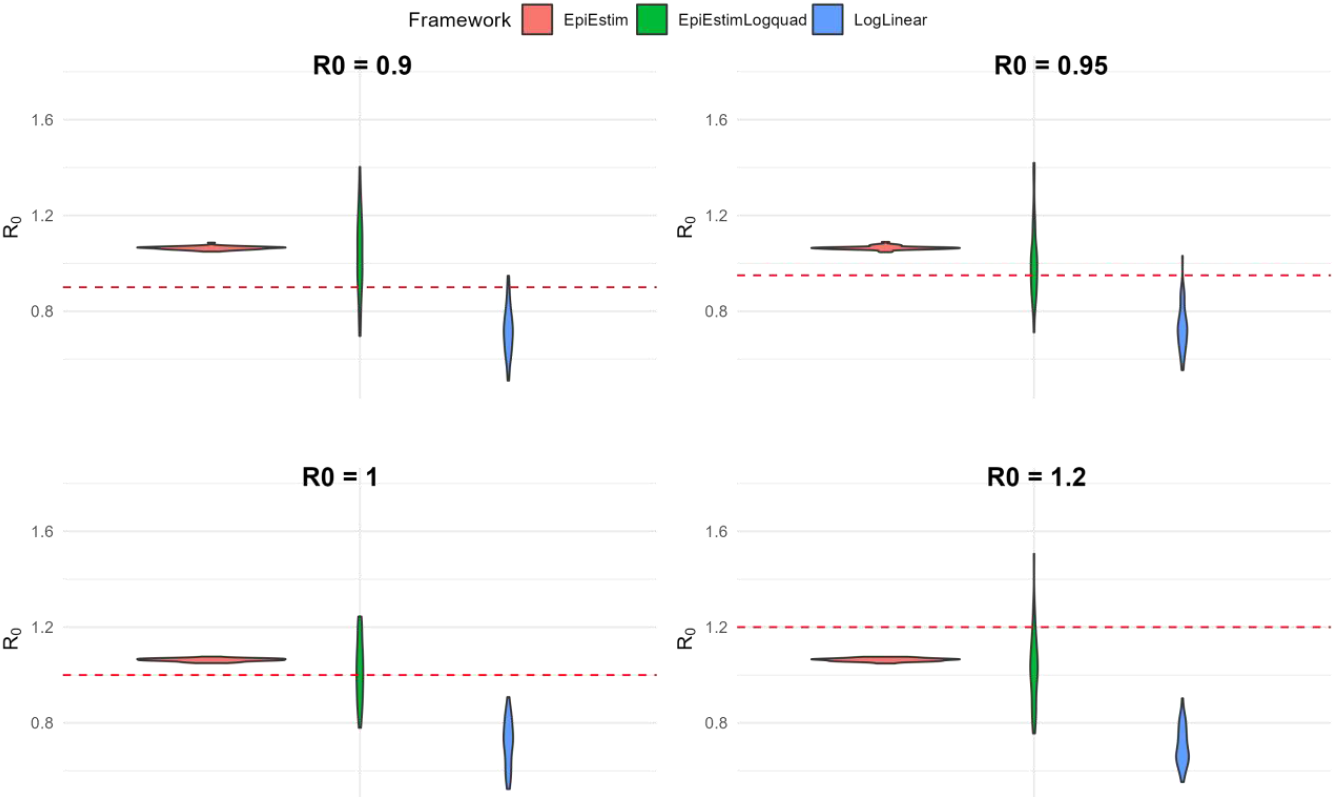
Distribution of average *R*_0_ estimates. Assuming a fixed reporting rate *λ*= 0.8 with 50 times simulation and population number 1e+8. Each sub-panel shows the distribution of the mean *R*_0_ estimates obtained for a given true *R*_0_ value around the kink (red dashed line). Abbreviations: LogLinear, growth rate follows log-linear distribution; EpiEstim, classic EpiEstim framework; EpiEstimLogquad, Log-quadratic adjustment of EpiEstim framework.

**Figure 3.**
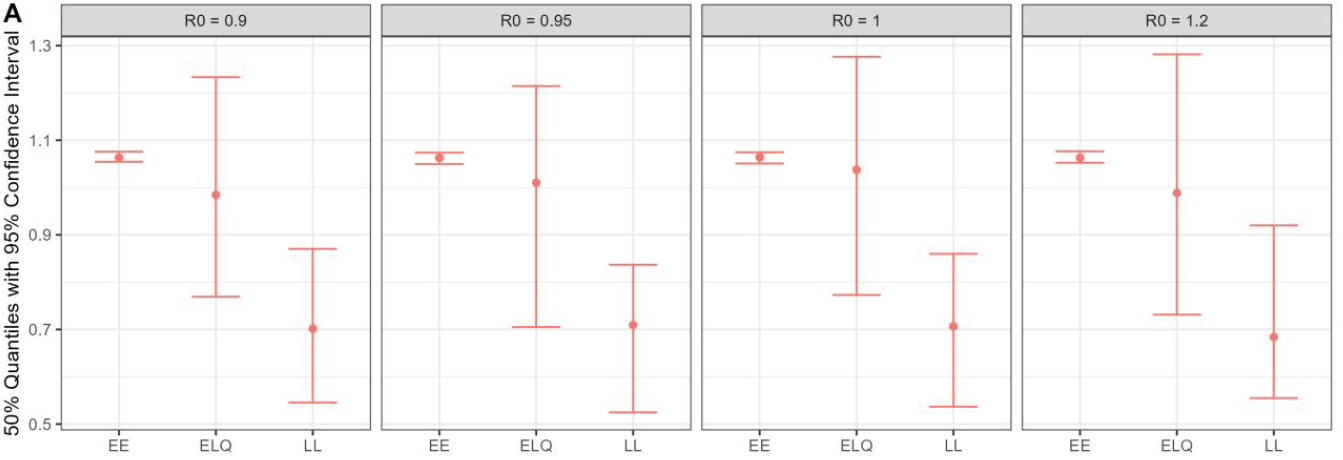
Estimation of 95% Confidence Interval corresponding to Figure 2. The generation interval of the mean is equal to 7 days and the standard deviation is 5 days. Method abbreviations: EpiEstim model (EE); EpiEstim Log-quadratic model (ELQ); Log-linear model (LL).

### Application to Outbreak Data

The performance of the proposed model has been examined over simulation data, in this section I applied it to the observation data of COVID-19 in practice reported by the John Hopkins Center for Systems Science and Engineering database focusing on four countries from June to August 2022 where kink trait was observed [15, 16]. These settings include Bulgaria, Japan, Poland, and South Korea, all of which experienced a substantial growth of cases around mid-June 2022 and a kink was observed. Different countries might not share the same reasons associated with the precipitous change. For each country, I fitted the incidence data with a log-quadratic regression model and then project the trajectory over the subsequent time window versus the EpiEstim model. I compared the trending difference of the resulting effective reproduction number by utilizing a sliding window of 3 days. For a varying range of sliding windows covering from 2 to 5 days, the qualitative outcome of estimates does not change (Supplementary material Figure S1-S6). The mean and standard deviation of the generation interval is 6 and 2 days respectively [17]. Figure 4 shows that the adjustment model generally led to upward trending of *R*_*t*_ for all four countries, generating higher values of *R*_*t*_ than their counterparts estimated through the traditional EpiEstim model. On the other hand, the estimates by EpiEstim are more irregular and zigzag-shaped, the estimates are curtailed to values below one within some specific intervals, deviating from the increasing spread of the pandemic. On average, *R*_*t*_ is increasing over time using the proposed model, and it is more unpredictable over time with the EpiEstim model.

**Figure 4.**
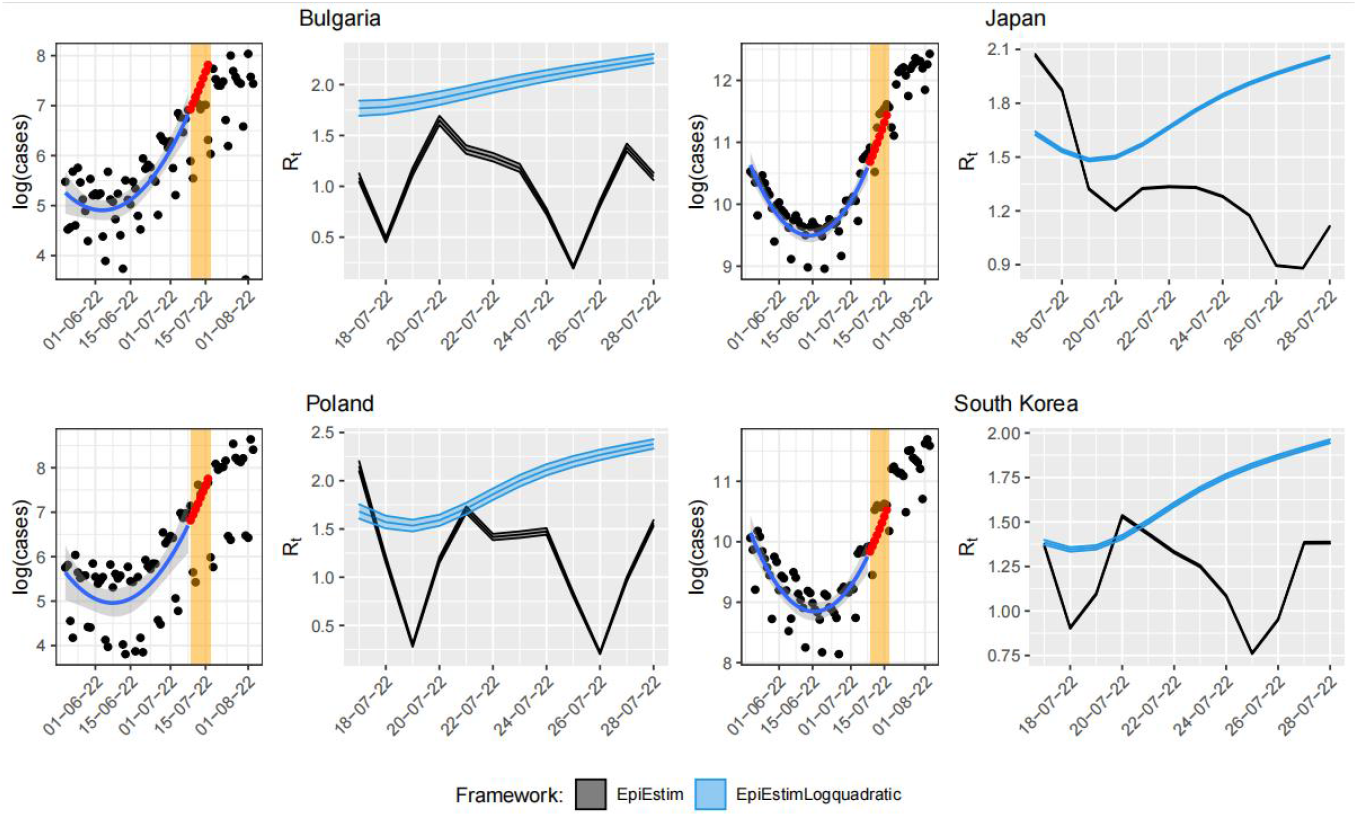
Comparison of *R*_*t*_ estimates. Obtained with (red and blue curves) and without (black points) adjustment of COVID-19 data in 4 countries: Bulgaria, Japan, Poland, and South Korea. Each sub-figure includes the log-transformation of the data and the regression line with confidence interval (left) and the *R*_*t*_ estimates one week ahead (right) obtained using a sliding window of 3 days and a generation interval of mean 6 days and standard deviation of 2 days. Abbreviations: LogLinear, growth rate follows log-linear distribution; EpiEstim, classic EpiEstim framework; EpiEstimLogquadratic, Log-quadratic adjustment of EpiEstim framework.

The violin plots corresponding to Figure 4 are illustrated in Figure 5. It can be seen that the *R*_*t*_ estimates approximately distribute in a range from 1.8 to 2.3 using the proposed model versus another range from 0.2 to 1.7 using the traditional EpiEstim model for Bulgaria. Near the kink, the former framework obtained a greater magnitude of estimation. These ranges are roughly (1.5, 2.1) versus (0.9, 2.1) for Japan, (1.5, 2.8) versus (0.2,2.3) for Poland, and (1.3, 1.9) versus (0.8, 1.5) for South Korea respectively. All countries attain similar trending of discrepancy. Generally, when more contagious variants of virus emerge and thus kink is present, the spread of the disease advances rapidly and the estimate of *R*_*t*_ is greater on average using the proposed framework versus classic models. The detection could potentially resemble multiple influencing drivers rather than a specific impacting factor.

**Figure 5.**
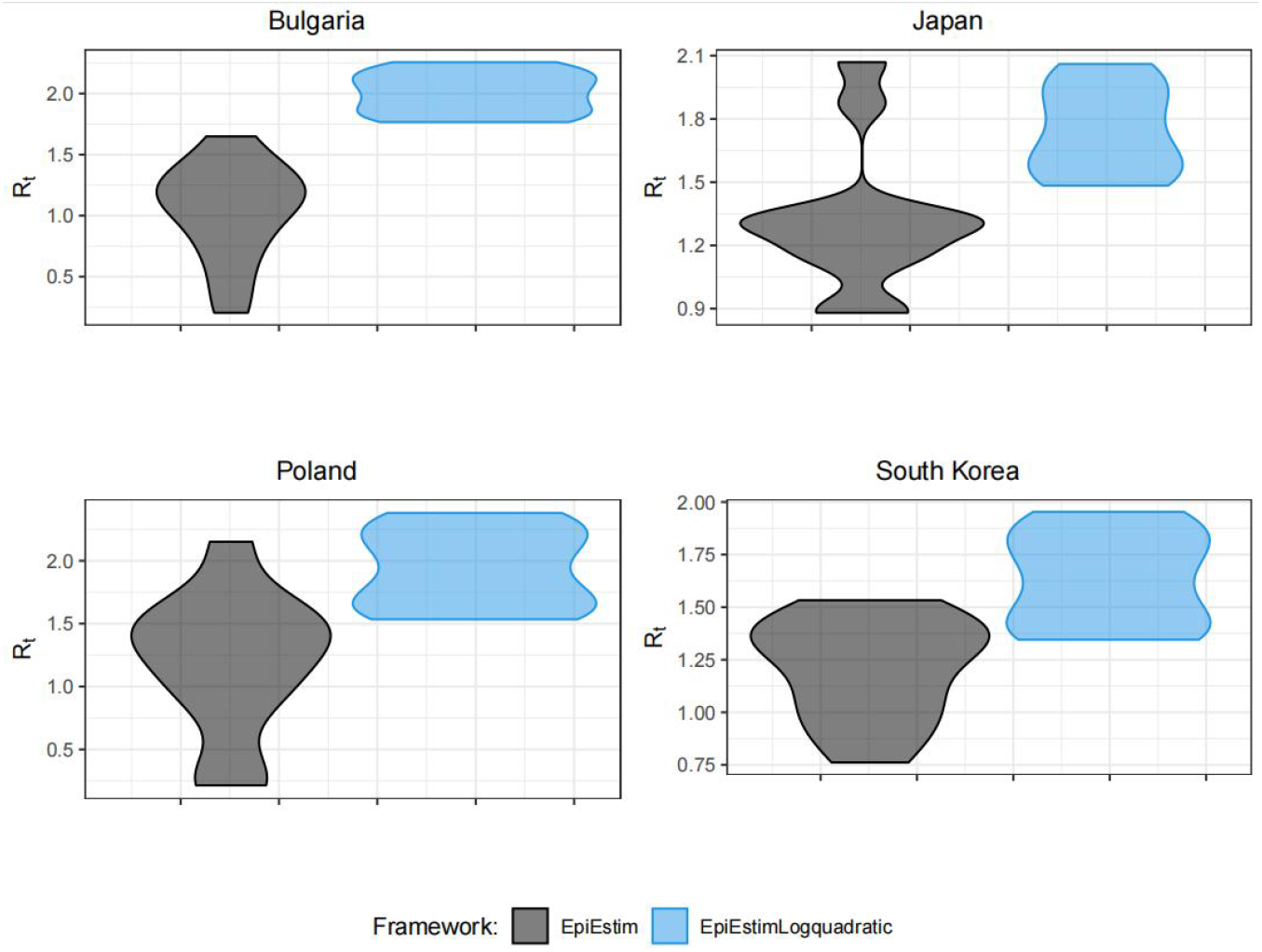
Violin plots corresponding to Figure 4. *R*_*t*_ estimates obtained using a sliding window of 3 days and a generation interval mean of 6 days and standard deviation of 2 days. Abbreviations: EpiEstim, classic EpiEstim framework; EpiEstimLogquadratic, Log-quadratic adjustment of EpiEstim framework.

## Discussions

I conceived an updated basic framework for the systematic bias of *R*_0_ that occurred in the early-variant transition stages of a shrinking epidemic accompanied by emergent variants, where kink traits were observed, when using traditional EpiEstim and log-linear methods accounting for transmissibility, data distribution, reporting rates, sliding windows, and generation intervals. I utilized the forward-imputation procedure to asymptotically project the outbreak one week ahead in association with a log-quadratic model to improve the accuracy estimates of the 95% confidence interval in four countries including Bulgaria, Japan, Poland, and South Korea from June to August 2022. The proposed correction aims to account for incorrect or missing reporting of infections applicable in settings where substantial sub-clinical infections, vaccine ineffectiveness, vaccination hesitancy, and unpreparedness of the population and healthcare system are observed [21]. In practice, the adjustment is meaningful in that I shed light on the necessity of updating countermeasures to infectious disease during its early-mutation stages potentially accompanied by vibrant variants as a great deal of asymptomatic or pre-symptomatic transmission advances and vaccines are not effective [20-28]. The correction may also apply to diseases characterized by abrupt alteration of the trajectory where more infectious variants of virus show up. In this study, I demonstrated the applicability of the revision to the classic EpiEstim and Log-linear method by curtailing the bias on the estimation of basic and effective reproduction numbers [24-30]. The update integrates the traits of EpiEstim and the log-linear frameworks, applying to the kink at the early-variant phases of transition. In particular, the bias observed in traditional package estimates is reduced and coverage is improved by the correction model around the kink. The adjusted estimates of *R*_0_ outperformed the estimates obtained with EpiEstim and log-linear methods in terms of estimate bias and produced improved coverage of 95% confidence intervals. This is partially attributable to the adjustment of the model closely complying with the principal traits of the real trajectory which is used for the forward imputation of the subsequent infections. I found a consistent upward trend of effective reproduction number close to the kink corner when more infectious variants emerged and vaccines potentially were not effective in four settings from June to August 2022.

Critically, the results are not sensitive to the choice of EpiEstim parameters, such as the interval of generation and the interval of the sliding window (Supplementary Figures S1–S9). This suggests that our adjustment has a much smaller effect when taking into account the potential co-founders. To evaluate the sensitivity of the estimates to the log-quadratic assumption, I test the performance of the method using a variety level of parameters. The results of this sensitivity analysis illustrated in the Supplementary Documentary indicate that the corrected model performs well near the kink accompanied by infectious variants of the pathogen.

The adjusted model reduced the bias estimates obtained with traditional EpiEstim and log-linear methods. The overestimation of reproduction number when *R*_0_ < 1 using EpiEstim reinforced the other findings in [20,22-29]. Additionally, I found that EpiEstim underestimated the reproduction number when *R*_0_ > 1 near the kink. This may partially be attributable to the reason that the transmission at the kink corner is rapid and the generation time is mostly shorter than usual.

### Limitations

Currently, COVID-19 has not yet been globally eliminated and experienced multiple waves of variants [31-39]. The proposed method partially incorporated the uncertainty of imputed cases, the heterogeneity of transmission dynamics, and the uncertainty of the growth rate estimates through the channels of sliding windows, data distribution, generation intervals, and reporting rates. The novel model also partially took into account the impact of estimate bias in terms of underestimation and overestimation. Estimation of the effective reproduction number can yield different outcomes using diverse frameworks accounting for systematic over- or under-estimation bias [40-43].

However, multiple limitations should be noted. First, I did not incorporate the uncertainty of other observed and unobserved factors potentially driving the kink transition such as changes in individual behaviors, contact patterns of the population, prior records of infections, non-compulsory and compulsory interventions, and their interactions [20, 25-33]. Studies found that non-compulsory measures reduced population mobility using mobile-phone data in Tokyo [8,34]. The survey conducted in Bulgaria implied that prior records of infections could confer certain levels of immunity protection, implicitly yielding an impact on the trajectory of later reinfection [35]. Second, other exterior driving factors may be playing roles as well, such as policies, environmental scenarios, urban space features, and potential interactions [36]. Third, I investigated the wrap-up effects resulting from the transition of emerging variants, however, I did not differentiate the effect of one driving factor from another. Neither did I differentiate the factor of variants from other drivers, nor the magnitude of each [36,38-39]. To accurately identify the stratified solution of the mathematical models for an outbreak incorporating these factors, more detailed measures and observations of relative metrics are needed (e.g., data on stratified distribution of urban space features including diversity of infrastructure buildings, environment scenarios including humidity, temperature, and other atmospheric conditions). The direct and accurate measure of the impact of health policies sometimes could be challenging and they might need time to take effect [36-40]. As a potential alternative, changes in population mobility, contact patterns, and other activities likely resulting from policies are discussed [36-42]. These refined measures can be in combination with graph analysis to capture the diversity in the urban space and environmental scenarios [36-41].

Future work can contribute to uncovering the association between the stratified uncertainty and the resulting parameter estimates as well as the impact of other potential driving factors when more information unfolds [36].

## Conclusions

The estimates using the proposed framework unveiled the consistent upward trending of effective reproduction numbers when the epidemic was transiting from convergence to divergence in four countries including Bulgaria, Japan, Poland, and South Korea from June to August 2022. The outcome implies that our log-quadratic and forward-imputation framework can yield improved precision of 95% confidence interval coverage with constraint or incorrect information and reduced the estimates of bias, which may prove meaningful in emerging novel epidemics, in which no effective vaccines are readily available. Although studies reported the estimates of virus variants, the non-linear transformation of the model especially the kink traits was rarely investigated [23,29-33], which impeded further analysis. The findings suggest that caution needs to be exercised when transmission of disease starts to slow down as it could be the starting point of other novel variants [30-33]. Hence, close monitoring of social measures is essentially needed during the outbreak [31,32].

I employed a log-quadratic trending model to infer incorrect or missing information approximately one week ahead in connection with in-equilibrium when considerable alteration of transmission occurs. The proposed framework improved the accuracy of range estimates of reproduction numbers in comparison with traditional statistical methods adjacent to the kink. Our analysis suggested the necessity of updating models to reduce the initial bias in reproduction number estimates during the occurrence of novel emerging variants when the estimates of principal parameters are critical for the calibration of public countermeasures at the turning corners [31-33,36].

## Data Availability

Supplementary materials are available online. The posted materials are not copyedited and are the sole responsibility of the authors, questions or comments should be addressed to the corresponding author. The R codes to reproduce this analysis are available upon correspondence.

## Author Contributions

I.N. conceptualized the study, designed models and implemented simulations, performed analyses, interpreted results, and wrote and revised the manuscript. I.N. reviewed and contributed to the revision of the manuscript.

## Acknowledgments

This work was supported by the China Ministry of Education Industry and Education Cooperation Project [grant no. 202002041005], Undergraduate Teaching Reform Foundation of Fujian University and Technology [grant no. 2022JG041], Graduate Teaching Reform and Textbook Publication Foundation of Fujian University and Technology [grant no. YJC22-1], National Social Science Foundation of China (22BGL007), Natural Science Funding of Fujian Province [grant no. 2020J01892], Fujian Zhi-lian-yun Supply Chain Technology and Economy Integration Service Platform, Fujian-Kenya Silk Road Cloud Joint R&D Center (2021D021), Fujian Provincial Department of Science and Technology, the Fujian Social Sciences Federation Planning Project (FJ2021Z006), and General program of Fujian Natural Science Foundation (2022J01941), Fujian Provincial Department of Education Project (No.JAT220230), Initial Scientific Research Fund in Fujian University of Technology (No.GY-Z220292), Fujian Provincial Department of Education Project (No. JAT220230), Research Project of Major Education and Teaching Reform in Fujian Universities (FBJG20190174).

## Data Availability

The outbreak data presented in this study are publicly available and are accessible from github.com/CSSEGISandData/COVID-19. The simulation data used in the study are available upon correspondence.

