## supplementary file for "Estimation of Near-kink Reproduction Numbers During the Emergent Variants of the COVID-19 Pandemic: Log-quadratic and Forward-imputation Approach"

### Robustness Test on Sliding Window

We performed a series of robustness checks to appraise the impact of the interval of the sliding window on the estimates of reproduction number attained with classic EpiEstim and the proposed framework. We used the simulation data sketched in the main manuscript and devised three potential types of sliding windows accounting for the vibrant change of transmission during the occurrence of variants. The sliding window of Figure S1, S3, and S5 is equal to 3, 4, and 5 days respectively.

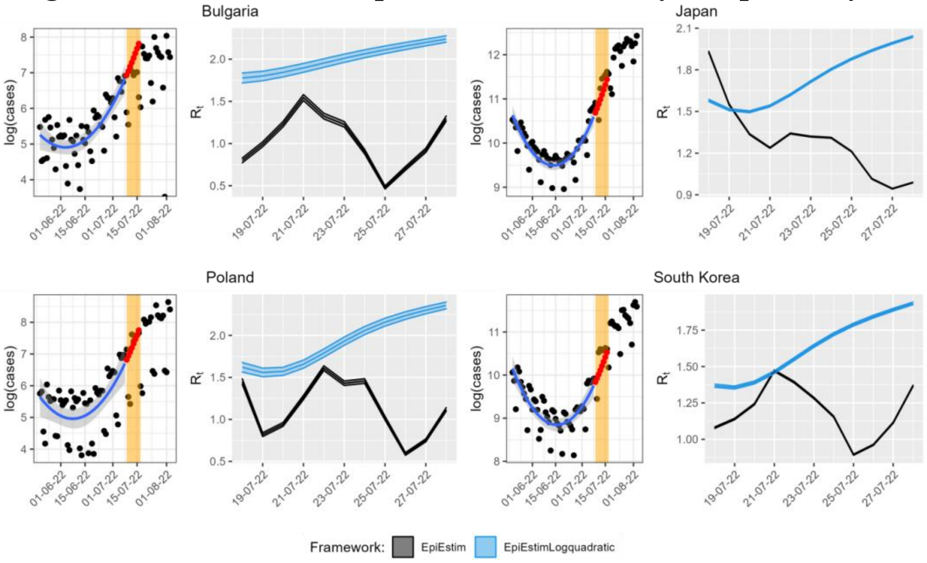


**Figure S1. Robustness test on sliding window.** Each subfigure includes the logarithm transformation of the data and the log-quadratic fitting curve with confidence interval (left blue), the $R_t$ estimates obtained using a sliding window of 3 days and a generation interval of mean 6 days and standard deviation of 2 days. The forecast of the subsequent one-week infections is delineated in red points and the orange rectangles denote the corresponding time window of prediction. Abbreviations: EpiEstim, classic EpiEstim framework; EpiEstimLogquadratic, Log-quadratic adjustment of classic EpiEstim framework.

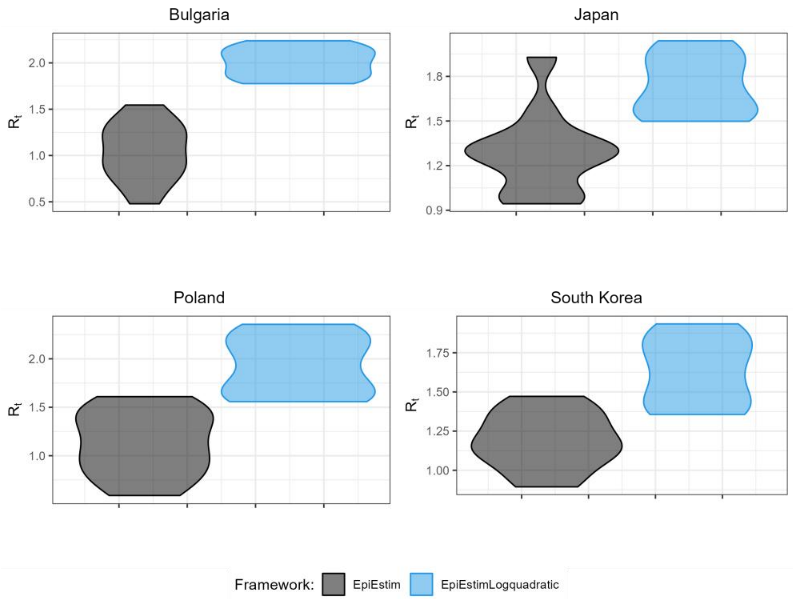

**Figure S2.** Violin plots corresponding to Figure S1 for Bulgaria, Japan, Poland, and South Korea. Abbreviations: EpiEstim, classic EpiEstim framework; EpiEstimLogquadratic, Log-quadratic adjustment of classic EpiEstim framework.

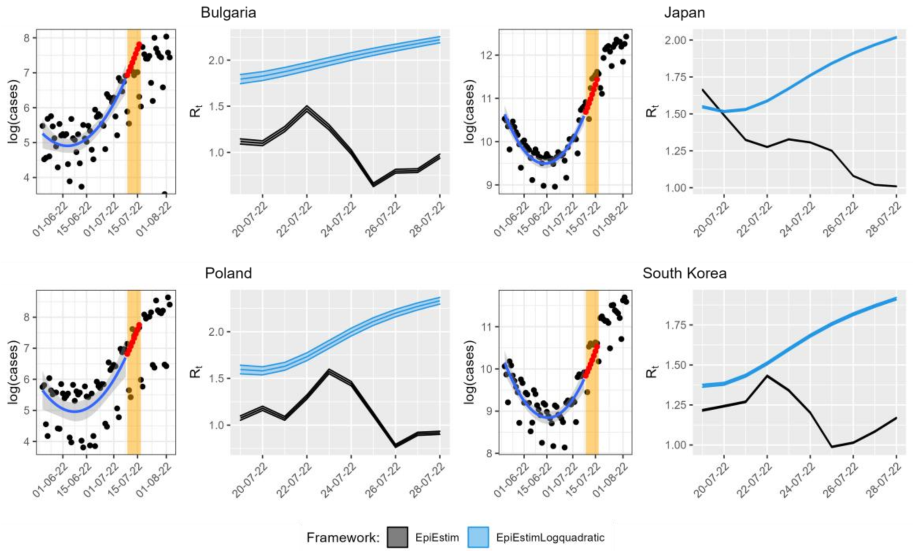


**Figure S3. Robustness test on sliding window.** Each subfigure includes the logarithm transformation of the data and the log-quadratic fitting curve with confidence interval (left blue), the $R_t$ estimates obtained using a sliding window of 4 days and a generation interval of mean 6 days and standard deviation of 2 days. The forecast of the subsequent one-week infections is delineated in red points and the orange rectangles denote the corresponding time window of prediction. Abbreviations: EpiEstim, classic EpiEstim framework; EpiEstimLogquadratic, Log-quadratic adjustment of classic EpiEstim framework.

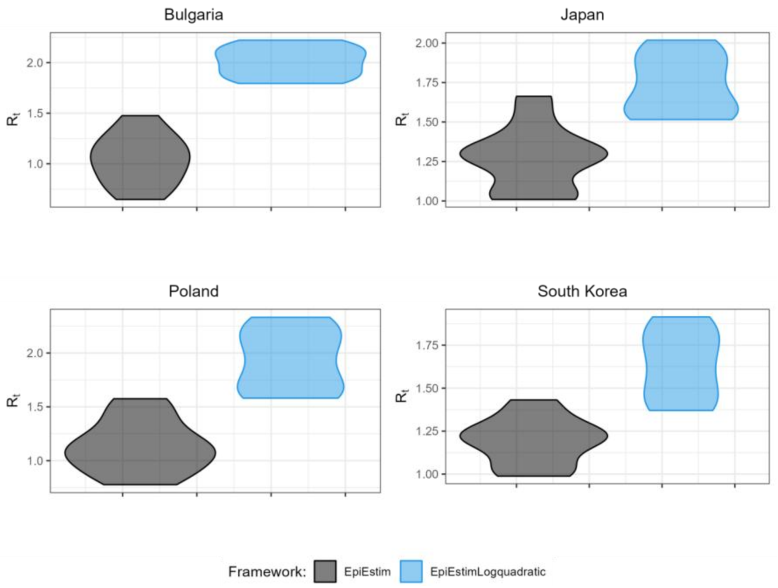


**Figure S4.** Violin plots corresponding to Figure S3 for Bulgaria, Japan, Poland, and South Korea. Abbreviations: EpiEstim, classic EpiEstim framework; EpiEstimLogquadratic, Log-quadratic adjustment of classic EpiEstim framework.

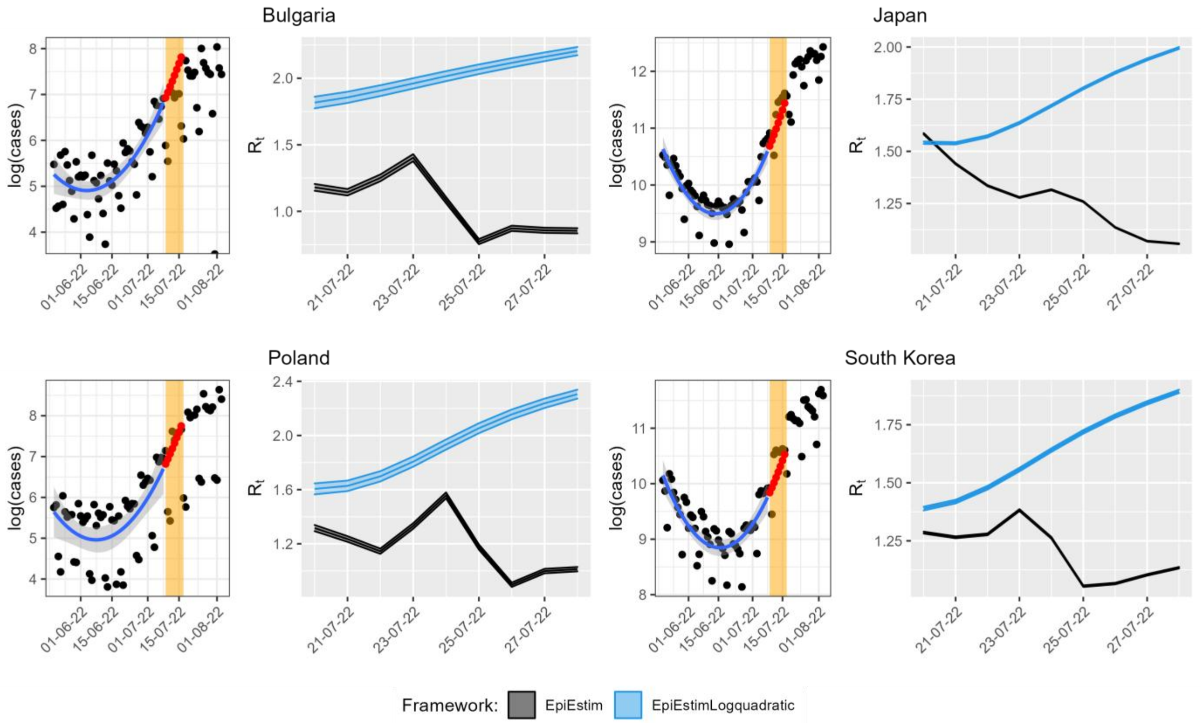


**Figure S5. Robustness test on sliding window.** Each subfigure includes the logarithm transformation of the data and the log-quadratic fitting curve with confidence interval (left blue), the $R_t$ estimates obtained using a sliding window of 5 days and a generation interval of mean 6 days and standard deviation of 2 days. The forecast of the subsequent one-week infections is delineated in red points and the orange rectangles denote the corresponding time window of prediction. Abbreviations: EpiEstim, classic EpiEstim framework; EpiEstimLogquadratic, Log-quadratic adjustment of classic EpiEstim framework.

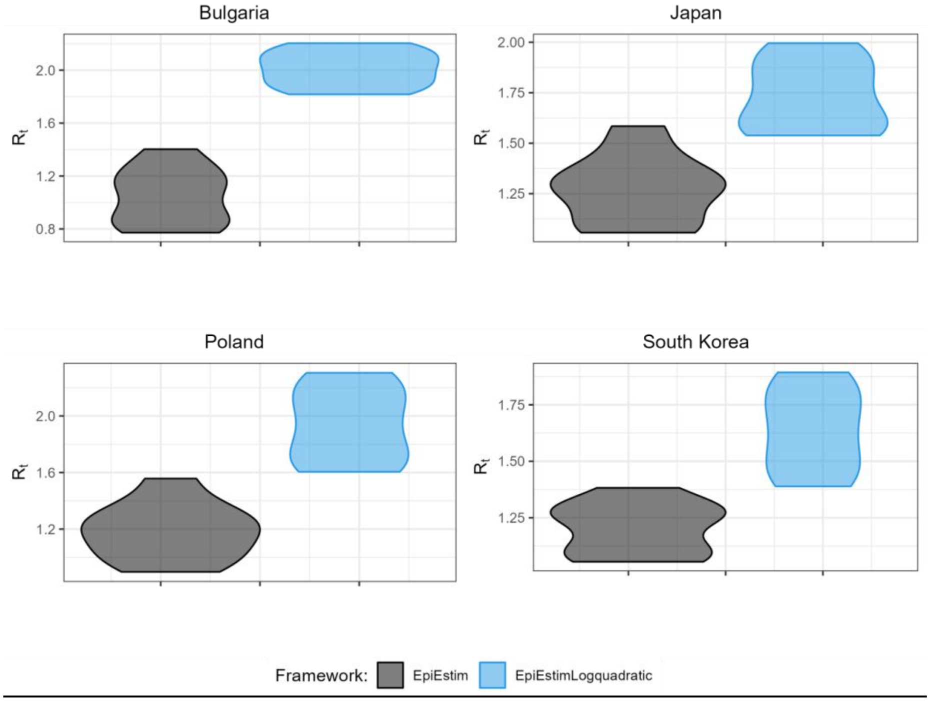

**Figure S6.** Violin plots corresponding to Figure S5 for Bulgaria, Japan, Poland, and South Korea.Abbreviations: EpiEstim, classic EpiEstim framework; EpiEstimLogquadratic, Log-quadratic adjustment of classic EpiEstim framework.

### Robustness test on Generation Interval

We performed as well robustness check to assess the impact of generation interval on the reproduction number estimates. We utilized the simulation data outlined in the manuscript and conceived three potential mean distributions. Figure S7 denotes the scenario where the generation interval mean and standard deviation are equal to 7 and 3 days respectively. Figure S8 denotes the scenario where the generation interval mean and standard deviation are equal to 8 and 4 days respectively. Figure S9 denotes the scenario where the generation interval mean and standard deviation are equal to 9 and 5 days respectively.

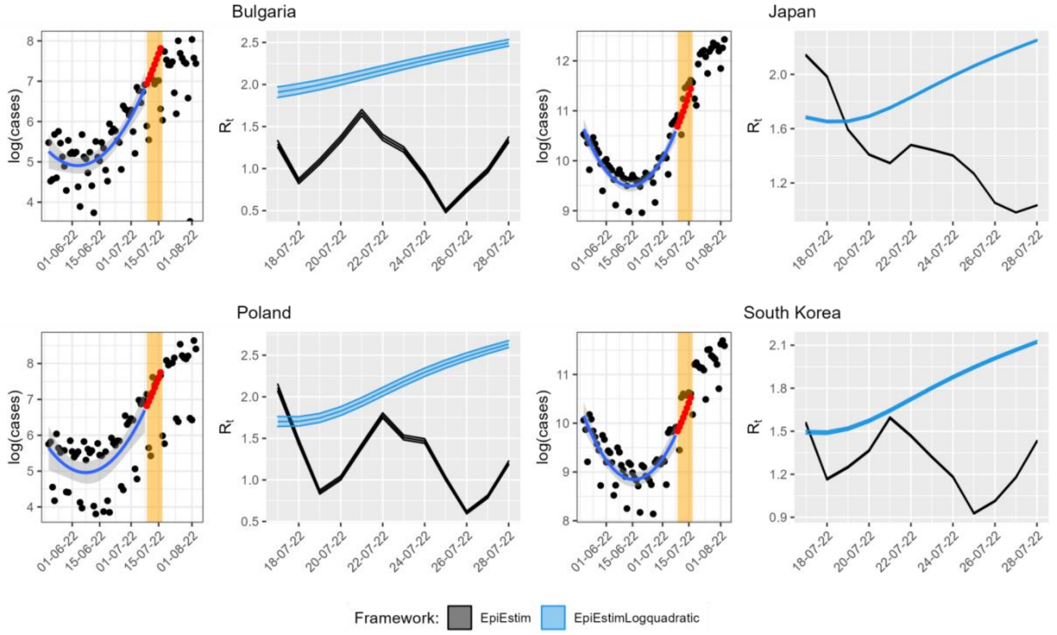


**Figure S7. Robustness check on generation interval.** Each subfigure shows the logarithm transformation of the data and the regression curve and the $R_t$ estimates obtained using a sliding window of 3 days and a generation interval mean of 7 days and standard deviation of 3 days. Abbreviations: EpiEstim, classic EpiEstim framework; EpiEstimLogquadratic, Log-quadratic adjustment of EpiEstim framework.

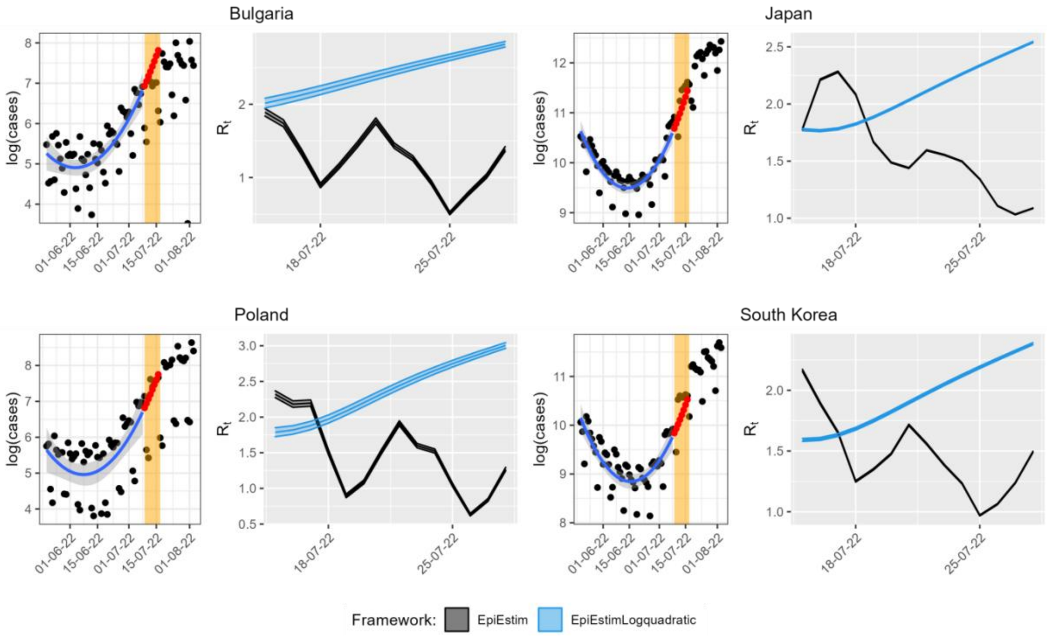


**Figure S8. Robustness check on generation interval.** Each subfigure shows the logarithm transformation of the data and the regression curve and the $R_t$ estimates obtained using a sliding window of 3 days and a generation interval of mean of 8 days and standard deviation of 4 days. Abbreviations: EpiEstim, classic EpiEstim framework; EpiEstimLogquadratic, Log-quadratic adjustment of EpiEstim framework.

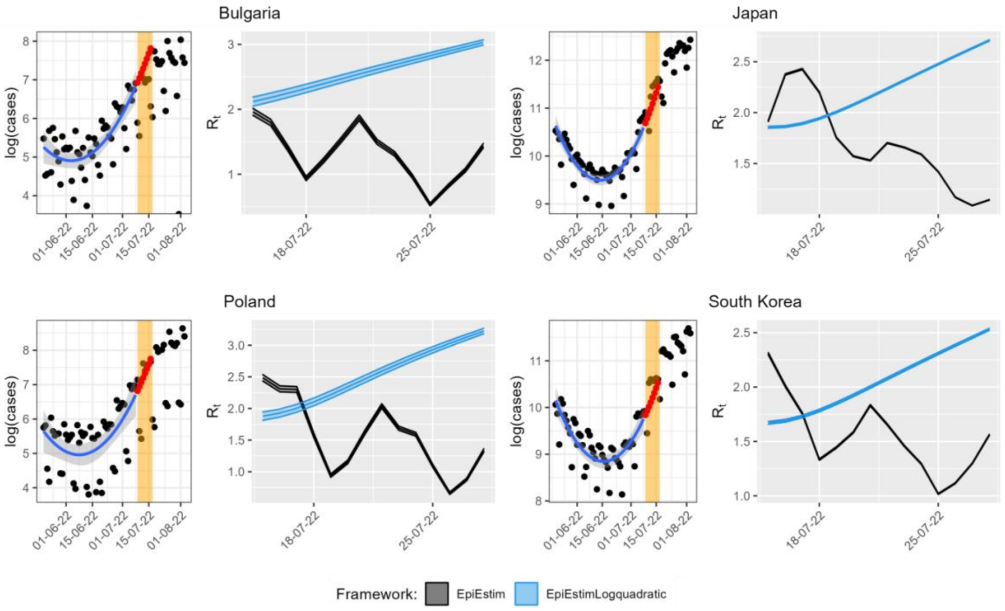


**Figure S9. Robustness check on generation interval.** Each subfigure shows the logarithm transformation of the data and the regression curve and the $R_t$ estimates obtained using a sliding window of 3 days and a generation interval mean of 9 days and standard deviation of 5 days. Abbreviations: EpiEstim, classic EpiEstim framework; EpiEstimLogquadratic, Log-quadratic adjustment of EpiEstim framework.

### Robustness test on reporting rate

We thereafter performed a robustness test to evaluate the impact of the varying reporting rates on the R0 estimates obtained in simulations. The results are shown in Figures S10-S17. The series of reporting rates take values from the set {0.2, 0.4,0.6,1.0}.

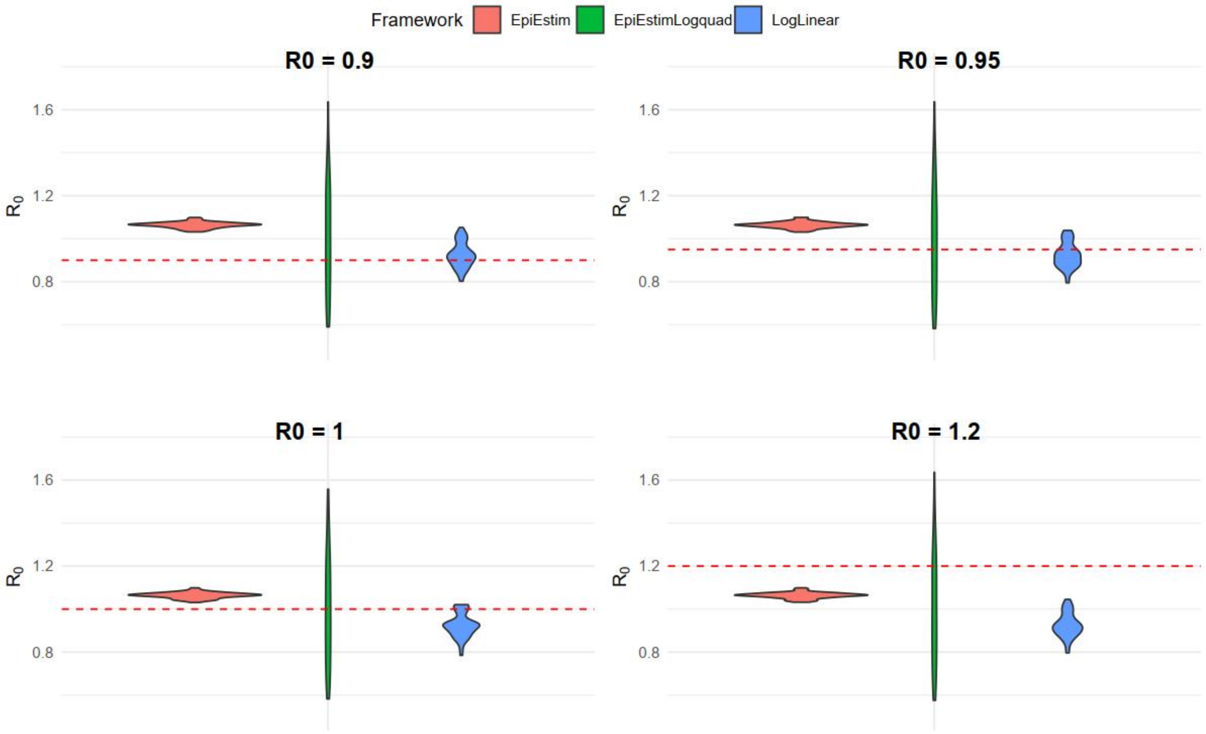


**Figure S10. Robustness test on reporting rates.** Suppose reporting rate $\lambda = 0.2$ . Each panel shows the distribution of the mean $R_0$ estimates obtained with 50 simulations for a true $R_0$ value (red dashed line). The generation interval of mean is equal to 7 days and the standard deviation is equal to 5 days. Abbreviations: EpiEstim, classic EpiEstim framework; EpiEstimLogquad, Log-quadratic adjustment of EpiEstim framework.

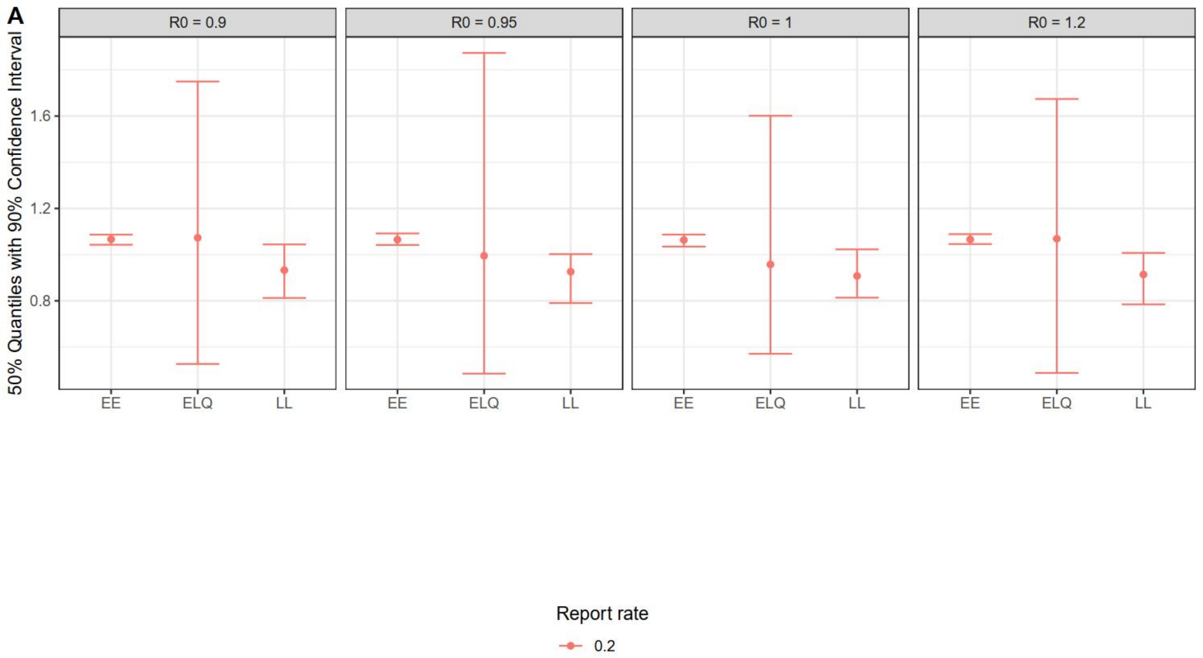


**Figure S11. 95% Confidence Interval corresponding to Figure S10.** Suppose reporting rate $\lambda = 0.2$. Each panel shows the distribution of the mean $R_0$ estimates obtained with 50 simulations for a true $R_0$ value (red dashed line). The generation interval mean is equal to 7 days and the standard deviation is equal to 5 days. Abbreviations: EE, EpiEstim model; ELQ, EpiEstimLogquadratic model; LL, Log-linear model.

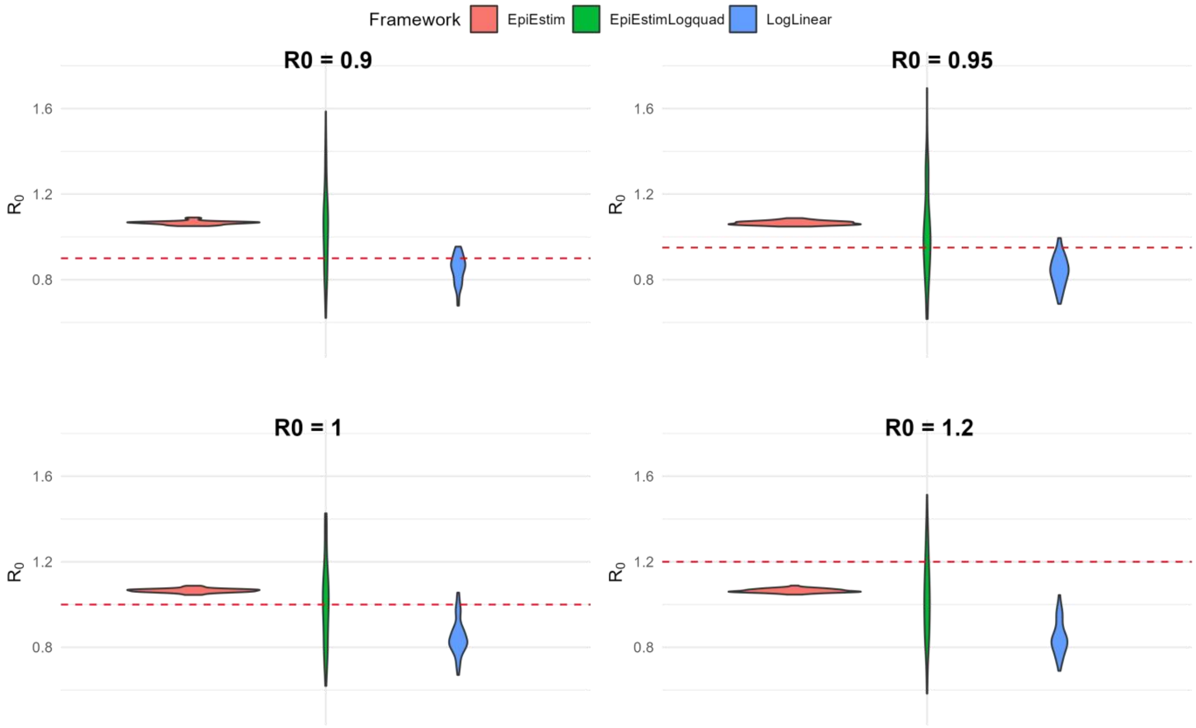


**Figure S12. Robustness test on reporting rates.** Suppose reporting rate $\lambda = 0.4$. Each panel shows the distribution of the mean $R_0$ estimates obtained with 50 simulations for a true $R_0$ value (red dashed line). The generation interval mean is equal to 7 days and the standard deviation is equal to 5 days. Abbreviations: EpiEstim, classic EpiEstim framework; EpiEstimLogquad, Log-quadratic adjustment of EpiEstim framework.

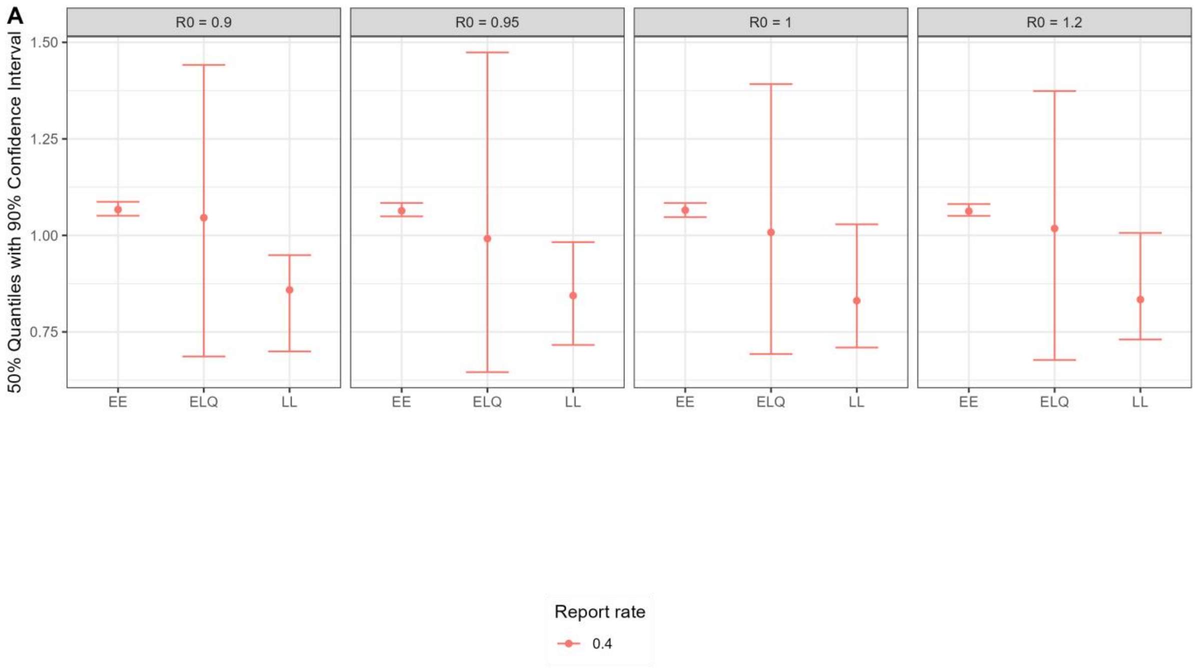


**Figure S13. 95% Confidence Interval corresponding to Figure S12. S**uppose reporting rate $\lambda = 0.4$ . Each panel shows the distribution of the mean $R_0$ estimates obtained with 50 simulations for a true $R_0$ value (red dashed line). The generation interval mean is equal to 7 days and the standard deviation is equal to 5 days. Abbreviations: EE, EpiEstim model; ELQ, EpiEstimLogquadratic model; LL, Log-linear model.

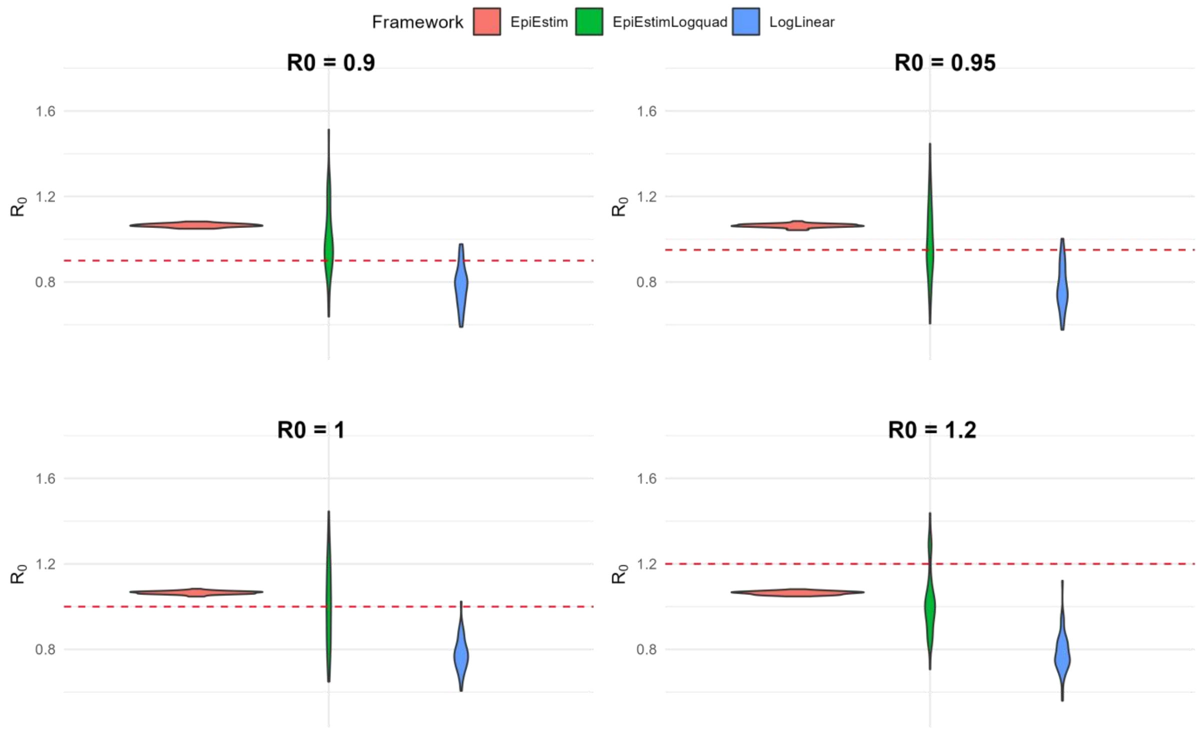


**Figure S14. Robustness test on reporting rates.** Suppose reporting rate $\lambda = 0.6$ . Each panel shows the distribution of the mean $R_0$ estimates obtained with 50 simulations for a true $R_0$ value (red dashed line). The generation interval of mean is equal to 7 days and the standard deviation is equal to 5 days. Abbreviations: EpiEstim, classic EpiEstim framework; EpiEstimLogquad, Log-quadratic adjustment of EpiEstim framework.

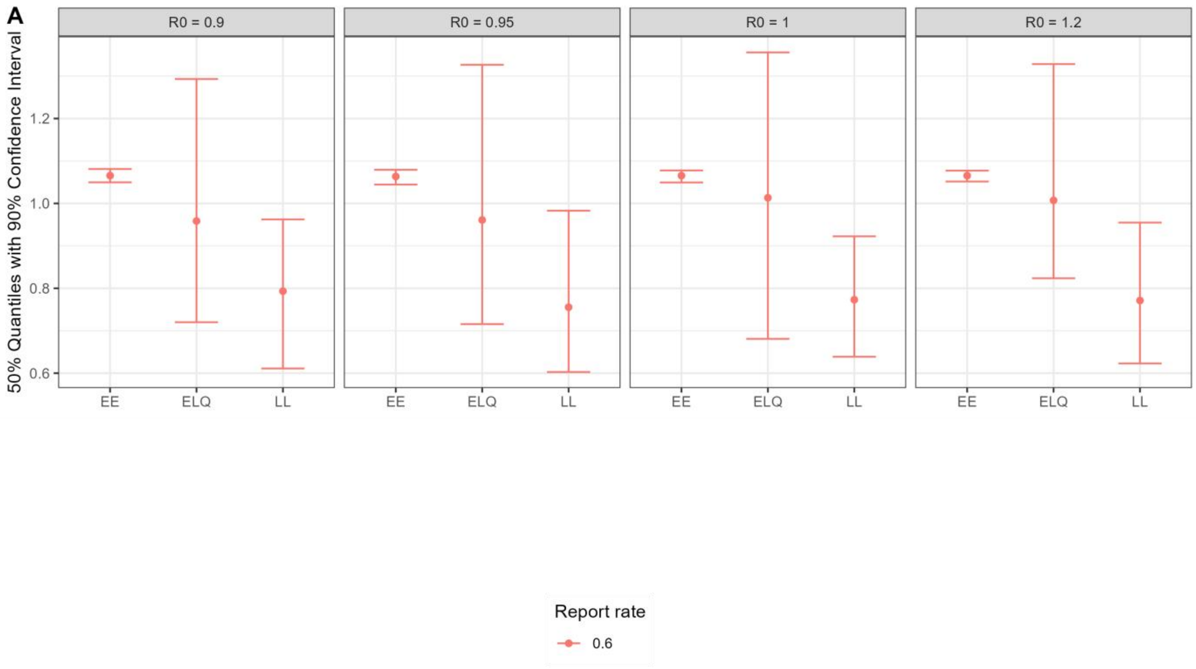


**Figure S15. 95% Confidence Interval corresponding to Figure S14.** Suppose reporting rate $\lambda = 0.6$ . Each panel shows the distribution of the mean $R_0$ estimates obtained with 50 simulations for a true $R_0$ value (red dashed line). The generation interval of mean is equal to 7 days and the standard deviation is equal to 5 days. Abbreviations: EE, EpiEstim model; ELQ, EpiEstimLogquadratic model; LL, Log-linear model.

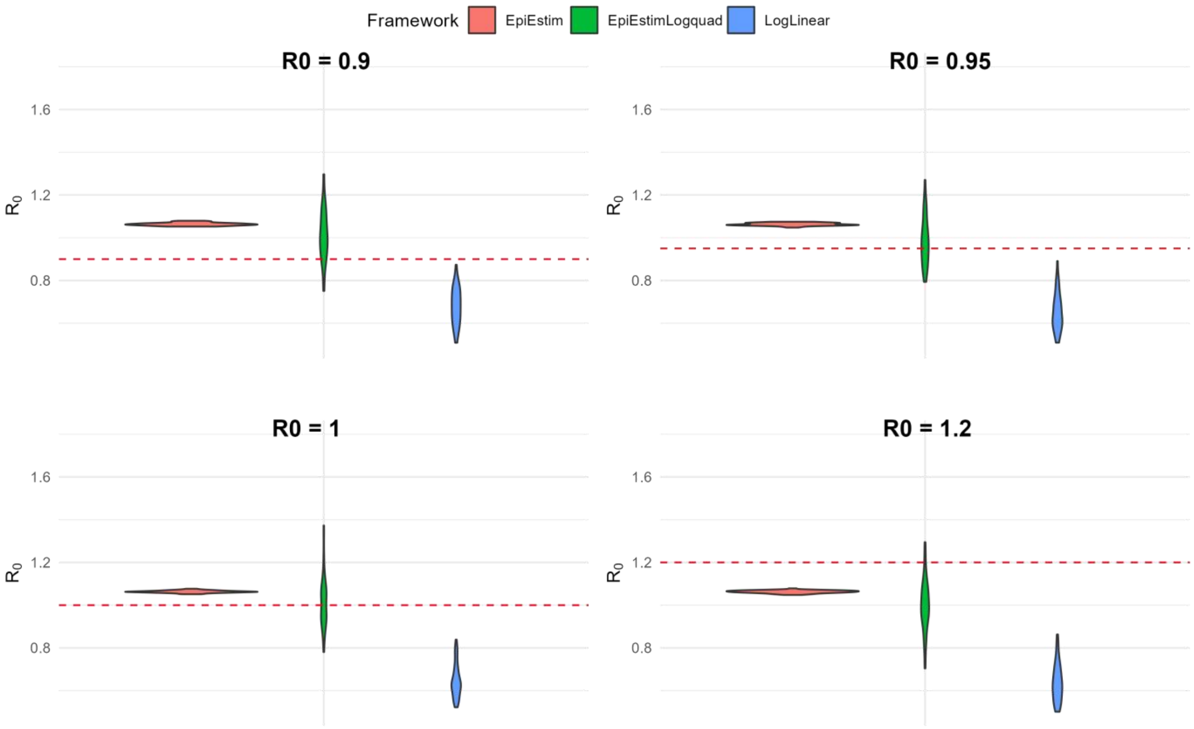


**Figure S16. Robustness test on reporting rates.** Suppose reporting rate $\lambda = 1.0$ . Each panel shows the distribution of the mean $R_0$ estimates obtained with 50 simulations for a true $R_0$ value (red dashed line). The generation interval of mean is equal to 7 days and the standard deviation is equal to 5 days. Abbreviations: EpiEstim, classic EpiEstim framework; EpiEstimLogquad, Log-quadratic adjustment of EpiEstim framework.

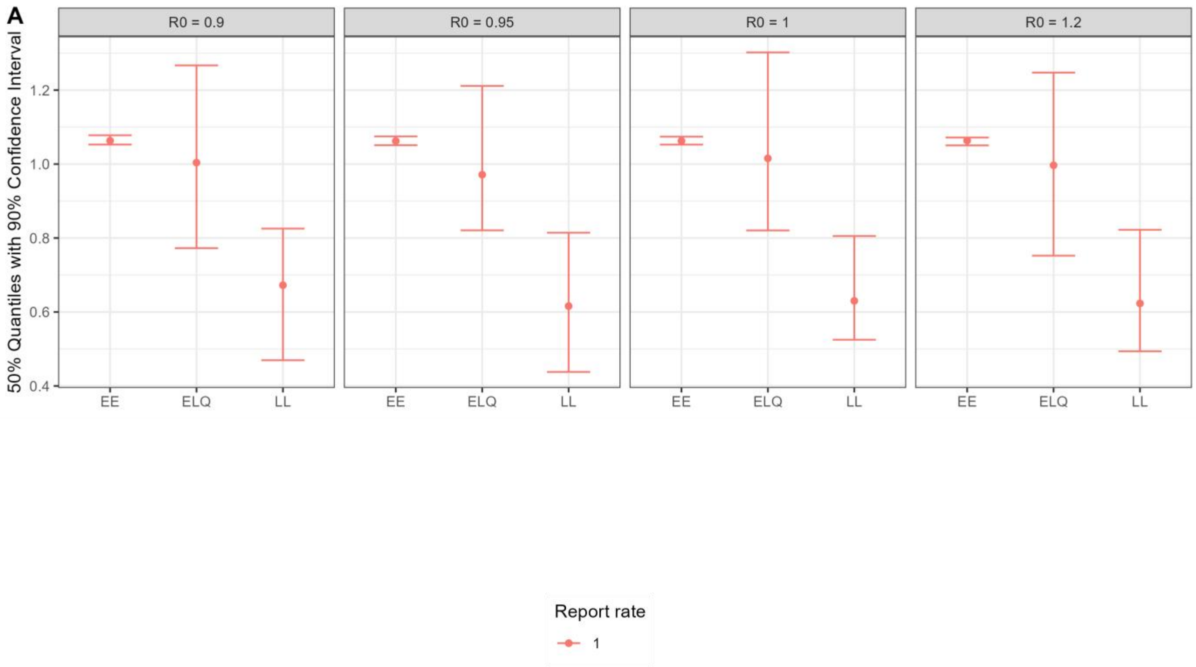


**Figure S17. 95% Confidence Interval corresponding to Figure S16.** Suppose reporting rate $\lambda = 1.0$ . Each panel shows the distribution of the mean $R_0$ estimates obtained with 50 simulations for a true $R_0$ value (red dashed line). The generation interval of mean is equal to 7 days and the standard deviation is equal to 5 days. Abbreviations: EE, EpiEstim model; ELQ, EpiEstimLogquadratic model; LL, Log-linear model.

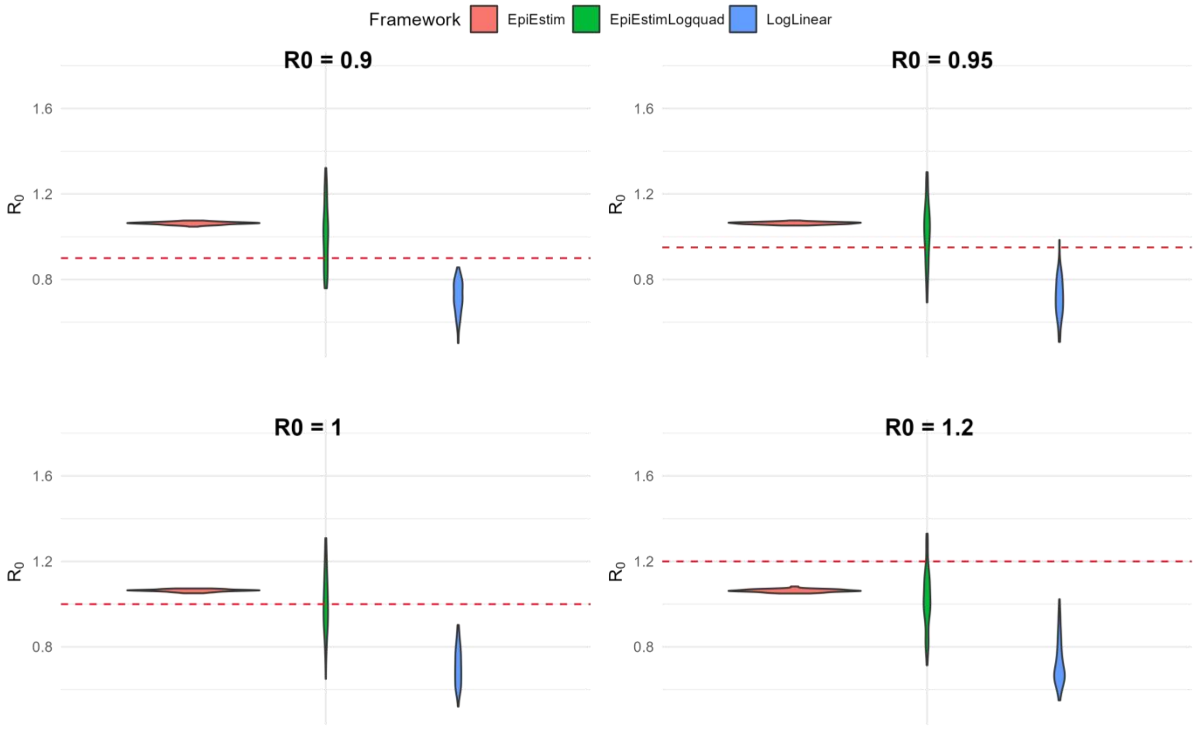

**Figure S18. Robustness test on reporting rates with Gamma Distribution.** Suppose reporting rate $\lambda = 0.8$ . Each panel shows the distribution of the mean $R_0$ estimates obtained with 50 simulations and population 1e+8 for a true $R_0$ value (red dashed line). The generation interval of mean is equal to 7 days and the standard deviation is equal to 5 days. Abbreviations: LogLinear, Log-linear framework; EpiEstim, classic EpiEstim framework; EpiEstimLogquad, Log-quadratic adjustment of EpiEstim framework.

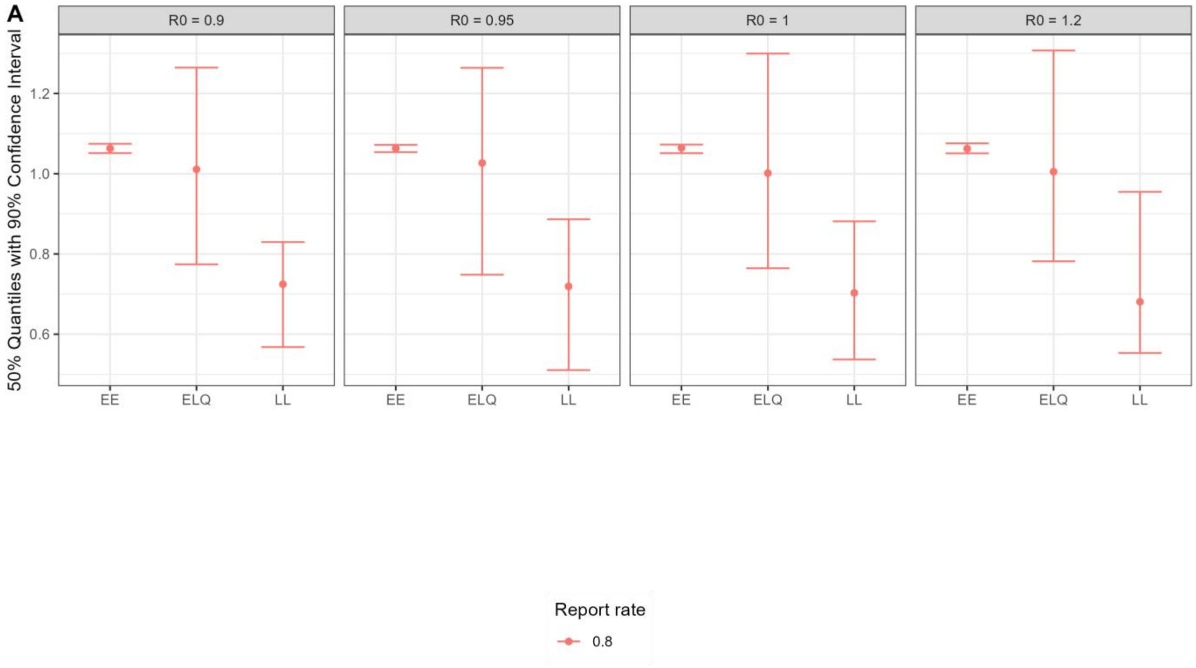


**Figure S19. Robustness test on reporting rates with Gamma Distribution corresponding to Figure S18.** Suppose reporting rate $\lambda = 0.8$ . Abbreviations: EE, EpiEstim model; ELQ, EpiEstimLogquadratic model; LL, Log-linear model.

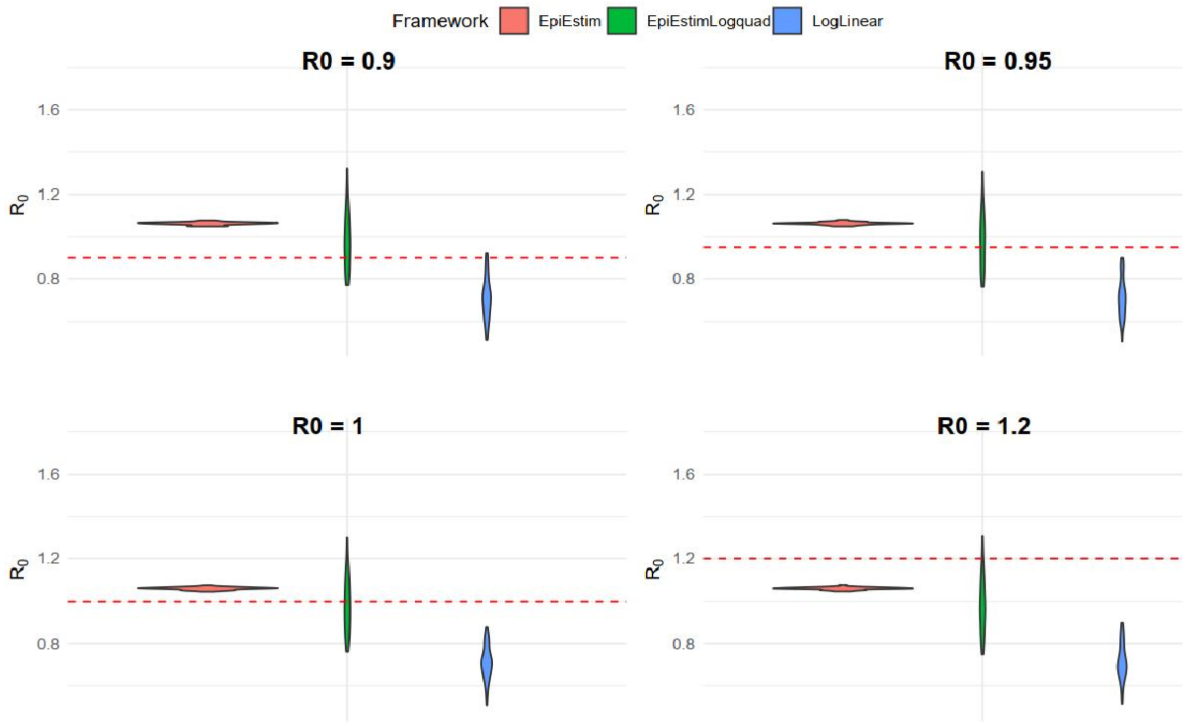


**Figure S20. Robustness test on reporting rates with Lognormal Distribution.** Suppose reporting rate $\lambda = 0.8$ . Each panel shows the distribution of the mean $R_0$ estimates obtained with 50 simulations and population 1e+8 for a true $R_0$ value (red dashed line). The generation interval of mean is equal to 7 days and the standard deviation is equal to 5 days. Abbreviations: LogLinear, Log-linear framework; EpiEstim, classic EpiEstim framework; EpiEstimLogquad, Log-quadratic adjustment of EpiEstim framework.

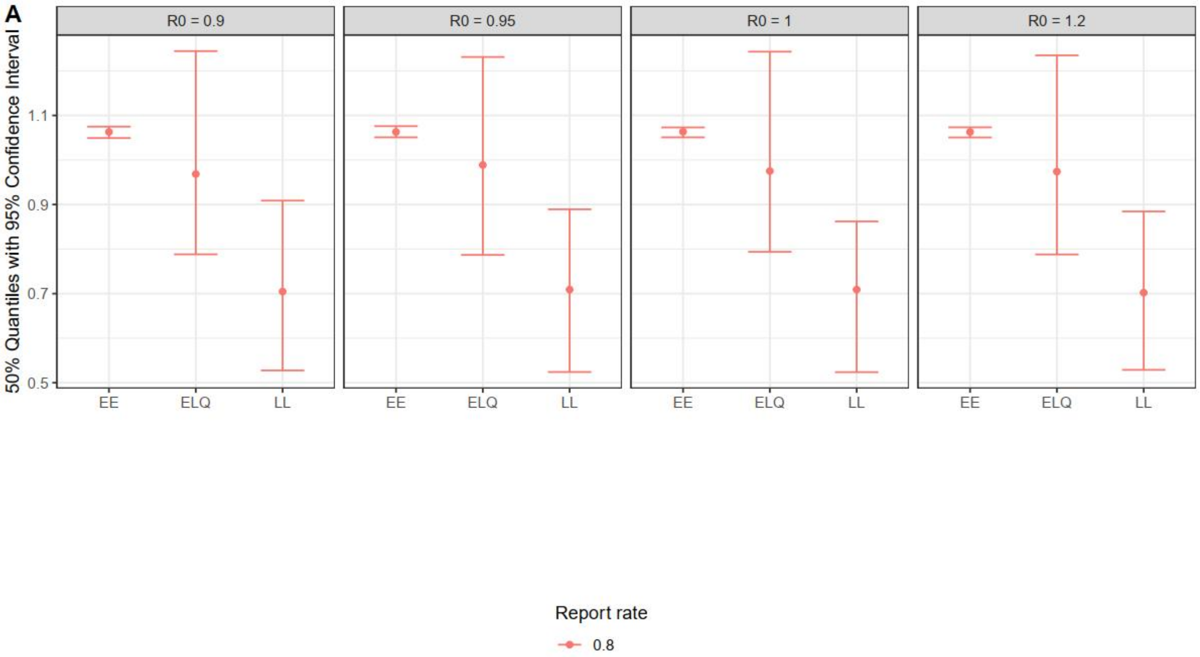


**Figure S21. Robustness test on reporting rates with Lognormal Distribution corresponding to Figure S20.** Suppose reporting rate $\lambda = 0.8$. Abbreviations: EE, EpiEstim model; ELQ, EpiEstimLogquadratic model; LL, Log-linear model.
